# Long-term motor and cognitive outcome of Deep Brain Stimulation in GBA-PD: the Italian PARKNET study

**DOI:** 10.1101/2024.12.23.24319546

**Authors:** Micol Avenali, Carlo A. Artusi, Roberto Cilia, Giulia Giannini, Giada Cuconato, Alberto Albanese, Nico G. Andreasi, Pietro Antenucci, Angelo Antonini, Laura Avanzino, Luca Baldelli, Anna R. Bentivoglio, Francesco Bove, Marco Bozzali, Giovanna Calandra-Buonaura, Ilaria Cani, Valerio Carelli, Francesco Cavallieri, Antoniangela Cocco, Filippo Cogiamanian, Fabiana Colucci, Pietro Cortelli, Alessandro De Biase, Francesca Di Biasio, Alessio Di Fonzo, Valentina D’Onofrio, Roberto Eleopra, Antonio E. Elia, Valentina Fioravanti, Danilo Genovese, Andrea Guerra, Alberto Imarisio, Claudia Ledda, Marco Liccari, Chiara Longo, Leonardo Lopiano, Maria C. Malaguti, Rachele Malito, Francesca Mameli, Silvia Marino, Raffaella Minardi, Pierfrancesco Mitrotti, Edoardo Monfrini, Claudio Pacchetti, Carla Piano, Vittorio Rispoli, Mario G. Rizzone, Luigi M. Romito, Luisa Sambati, Mariachiara Sensi, Chiara Sorbera, Francesca Spagnolo, Cristina Tassorelli, Francesca Valentino, Franco Valzania, Roberta Zangaglia, Maurizio Zibetti, the Italian PARKNET Study Group, Enza Maria Valente

## Abstract

Deep brain stimulation (DBS) is an established therapeutic option for Parkinson Disease (PD), with demonstrated efficacy on motor symptoms also in patients carrying *GBA1* variants (GBA-PD). However, it was recently suggested that DBS may accelerate cognitive decline in this frequent genetic subgroup, raising major concerns on its indication. Primary aim of this study was to investigate the possible additive effects of *GBA1* genotype and DBS implant on cognitive deterioration and other non-motor features in the long term. As secondary aims, we assessed the clinical outcomes of DBS-GBA-PD stratified by *GBA1* variant classes (severe/complex, mild, risk, unknown), and by different DBS targets (subthalamic nucleus or globus pallidus).

This is a multicenter retrospective, controlled, Italian cohort study involving 15 tertiary level Movement Disorder Centers contributing to the PARKNET cohort.

Demographic, motor, cognitive and other non-motor features were collected at baseline after 1, 3 and 5 years, between years 2005 and 2021. We selected 615 PD participants who either underwent DBS surgery (430 DBS-nonGBA-PD and 109 DBS-GBA-PD) or fulfilled the same criteria for DBS eligibility but eventually were not operated (76 nonDBS-GBA-PD).

Assessments included motor, cognitive and other non-motor features, as well as long-term complications, across all groups and time points. Within-group longitudinal outcome changes, between-group differences, and subgroups analyses stratified by *GBA1* variant classes and DBS targets were performed.

At baseline, the three cohorts were largely matched for demographic, motor, cognitive and other non-motor features. Longitudinally, both DBS groups showed marked improvements of motor symptoms and quality of life, a benefit which was absent in nonDBS-GBA-PD. Cognitive deterioration, as well as hallucinations and urinary problems, significantly increased in both GBA-PD groups compared to nonGBA-PD, regardless of DBS. No relevant differences in the clinical outcomes emerged upon stratification of GBA-PD for variant classes or for DBS targets, up to 3 years after surgery.

DBS represents a valid therapeutic option for GBA-PD, as it generates prolonged benefits on motor symptoms and quality of life while not modifying the occurrence of cognitive deterioration and other non-motor features.

## Introduction

Heterozygous *GBA1* variants represent the most common genetic risk factor for Parkinson Disease (PD), occurring in ∼10% patients worldwide.^1,2^ PD subjects carrying *GBA1* mutations (GBA-PD) are more likely to develop early motor complications and faster cognitive deterioration, and present a higher risk of dementia, compared to non-mutated patients (nonGBA-PD).^3–5^

Deep brain stimulation (DBS) is an established therapeutic option to improve motor symptoms in complicated phases of PD,^6,7^ also in cases with proven genetic etiology.^8,9^ Studies investigating DBS outcomes in GBA-PD have consistently shown marked motor improvements, with significant reduction of fluctuations, dyskinesias and dosage of dopaminergic medications.^10–13^ Yet, a large multicentre study reported a more rapid cognitive decline in GBA-PD subjects who underwent DBS of the subthalamic nuclei (STN), compared to both operated nonGBA-PD and non-operated GBA-PD. This suggested the occurrence of additive, detrimental effects of the *GBA1* genotype and DBS surgery on cognitive outcome,^13^ and raised serious concerns about offering DBS to GBA-PD individuals.

We recently assessed the outcome of DBS in two large cohorts of GBA-PD versus non-GBA-PD subjects, confirming a similar motor improvement but a more rapid cognitive deterioration of mutated individuals at 5-year follow-up.^14^ Analogous results were reported in a smaller cohort of Spanish patients.^15^ However, in both studies, the lack of a non-DBS GBA-PD cohort did not allow discriminating the relative impact of the genotype and the surgical procedure itself on the rate of cognitive decline.

Another unanswered issue relates to potential differences of DBS outcomes in carriers of different *GBA1* variants as, for instance, it is known that “severe” variants are associated with a worse cognitive outcome than other classes.^5,16–19^ Although a stratification of DBS-GBA-PD according to variant classes was attempted, subgroups were too small for statistical comparison, and no relevant conclusions could be drawn.

Finally, a single study on seven GBA-PD patients who underwent DBS either on the STN or the globus pallidus pars interna (GPi) suggested that the latter target could be associated with less cognitive worsening at 1-2 years follow-up, questioning what the optimal DBS target for GBA-PD could be.^20^

The present study aims to address the unsolved question about the possible additive effects of *GBA1* genotype and DBS implant on cognitive deterioration and other non-motor features in the long term. As secondary aims, we looked at possible differences in the clinical outcome of DBS-GBA-PD stratified for GBA1 variant classes (severe/complex, mild, risk, unknown), and by different DBS targets (subthalamic nucleus or globus pallidus).

## Materials and methods

This is a multicentre, retrospective, controlled, cohort study involving 15 tertiary level Movement Disorder Centres in the frame of the Italian PARKNET project (see Supplementary Methods). Ethics approval was obtained by the respective Committees and all patients provided written informed consent.

### Cohorts’ recruitment

In addition to the 365 patients included in our previous study (DBS-PD former cohort),^14^ we recruited:

1. a validation DBS cohort of 174 subjects (vDBS-PD) who underwent surgery according to eligibility criteria^21,22^ between years 2005-2021, fulfilling the following requirements:

− whole *GBA1* gene sequenced;^14^

− absence of pathogenic/likely pathogenic variants in other major PD-related genes;

− detailed clinical data available at pre-DBS and after 1, 3 and, when available, 5 years from surgery.

Based on *GBA1* genotype, subjects were stratified into vDBS-GBA-PD (n=36) and vDBS-nonGBA-PD (n=138).

1. 2) 76 non-operated GBA-PD subjects (nonDBS-GBA-PD), ascertained between years 2005-2021 and fulfilling the following criteria:

− heterozygosity for a *GBA1* variant

− absence of pathogenic/likely pathogenic variants in other major PD-related genes;

− an identifiable time in patient’s history (defined “baseline”) in which subjects met the same eligibility criteria for DBS surgery as the DBS cohort^21,22^ (Supplementary Table 1):

− detailed clinical data available at baseline and after 1, 3 and, when possible, 5 years.

Despite being eligible, these patients eventually did not undergo DBS for one or more of the following reasons: i) refused brain surgery (23.5 %); ii) opted for modification of routine oral therapy (70.3%); iii) opted for subcutaneous apomorphine therapy or intrajejunal levodopa/carbidopa (14.1 %); iv) lack of social/familiar support (6.2%); v) general major contraindication for surgery such as systemic comorbidities or parenchymal/structural brain lesions (12.5%).

*GBA1* variants were subclassified into 5 classes (mild, severe, complex, risk and unknown) as reported.^23,24^

### Clinical Assessment

Clinical data were retrospectively collected from records at four timepoints: baseline (T0) (for DBS: maximum 6 months before surgery; for nonDBS-GBA-PD: time of evaluation for surgical eligibility), after 1 (T1), 3 (T3) and, when available, 5 years (T5).

Mainly, we collected the following data: MDS-Unified Parkinson’s Disease Rating Scale part III (MDS-UPDRS-III) scores in ON and OFF medication phase (stim-ON after surgery),, Hoehn & Yahr (HY) stage, Mattis Dementia Rating Rating Scale (MDRS), presence of motor fluctuations, freezing of gait, autonomic symptoms, impulse control disorders (ICD), dementia, depression, hallucinations, inability to walk and recurrent falls.

A detailed list of the obtained demographic and clinical features is presented in Supplementary Methods.

### Statistical Analysis

Statistical analyses were performed using Statistical Package for the Social Sciences (SPSS v.26), with significance set at *p*<0.05 for all tests. The newly recruited DBS-PD validation cohorts (138 vDBS-nonGBA-PD and 36 vDBS-GBA-PD) were compared at each timepoint and longitudinally using the same methodology as reported in our former study,^14^ to ensure replicability.

Merged cohorts consisted of 430 DBS-nonGBA-PD and 109 DBS-GBA-PD patients, which were compared with the newly recruited group of 76 nonDBS-GBA-PD subjects (total number: 615 subjects).

Demographic characteristics among the three groups were assessed using Chi-squared for dichotomous variables, and Kruskal-Wallis’s test with Bonferroni *post hoc* correction for numerical variables. For each timepoint, comparisons among groups were performed using Chi-squared and logistic regression for categorical variables and Kruskal-Wallis’s test followed Bonferroni *post-hoc* adjustment for numerical variables. Longitudinal analysis was performed over 5 years using a linear mixed-effect model, with groups as independent variable; age, sex, years of education and disease duration were added as covariates. Statistical methodology used to assess differences in clinical outcomes of DBS-GBA-PD stratified by *GBA1* variant classes (severe/complex, mild, risk, unknown), and DBS targets (subthalamic nucleus or globus pallidus) is detailed in Supplementary Methods.

## Results

### Replication analysis in the DBS-PD validation cohort

The validation cohort included 174 participants who underwent bilateral DBS (138 vDBS-nonGBA-PD and 36 vDBS-GBA-PD). Frequency of *GBA1* variant classes was comparable to that of the former cohort (Supplementary Fig. 1).^14^

To assess homogeneity between former and validation cohorts, we first compared vDBS-GBA-PD versus vDBS-nonGBA-PD groups at each timepoint and longitudinally, using the same methodology as our previous study (Supplementary Results).^14^ This validation analysis generated fully consistent results, allowing us to merge the two cohorts to increase sample size and statistical power. We reached an overall numerosity of 430 DBS-nonGBA-PD and 109 DBS-GBA-PD subjects, which were then compared with the newly recruited group of 76 nonDBS-GBA-PD subjects. The following results refer to comparisons among these three groups, amounting to a total number of 615 subjects included in the present study. The list of *GBA1* variants detected in the entire cohort is reported in Supplementary Table 2.

### Baseline characteristics

At baseline, DBS-GBA-PD were slightly younger and had more dyskinesias and orthostatic hypotension symptoms than other groups. Moreover, both DBS groups had a slightly lower age at PD onset, longer disease duration, higher LEDD intake and more prevalent freezing of gait compared to non-operated subjects. All other motor and non-motor features, cognitive scores and quality of life (QoL) scores were comparable across the three groups (Table 1).

**Table 1.** Demographic characteristic at baseline.

|  | DBS-nonGBA-PD<br>(a)<br>n=430 | DBS-GBA-PD<br>(b)<br>n=109 | nonDBS-GBA-PD<br>(c)<br>n=76 | p | Post Hoc |
| --- | --- | --- | --- | --- | --- |
| Age at baseline (years) | 57.4 ± 7.7 | 53.5 ± 8.4 | 57.7 ± 8.1 | **< 0.001** | **b<a,c** |
| Age at PD onset (years) | 46.4 ± 8.2 | 44.2 ± 8.3 | 50.3 ± 7.5 | **< 0.001** | **c>a,b** |
| Age at DBS (years) | 57.7 ± 7.6 | 53.5 ± 8.4 | - | **< 0.001** | **b<a** |
| Sex (F/M) | 137/293 | 42/67 | 28/48 | ns | - |
| Disease duration at DBS (years) | 10.9 ± 4.4 | 9.1 ± 3.7 | 7.5 ± 3.0 | **< 0.001** | **b<a; c<a,b** |
| Education (years) | 11.8 ± 3.7 | 11.4 ± 3.8 | 12.5 ± 3.8 | ns | - |
| DBS target: STN vs GPi (% STN) | 92.1% | 89.4% | - | ns | - |
| Family history for PD (% yes) | 85/404 (21%) | 42/107 (39.3%) | 35/75 (46.7%) | **< 0.001** | **a<b,c** |
| Hoehn & Yahr | 2.2 ± 0.6 | 2.3 ± 0.6 | 2.1 ± 0.5 | ns | - |
| LEDD (mg) | 1053.58 ± 415.8 | 1095.5 ± 400.2 | 790.8 ± 297.2 | **< 0.001** | **c<a,b** |
| MDS-UPDRS-III (OFF med) | 41.6 ± 14.4 | 43.1 ± 16.3 | 36.2 ± 13.4 | ns | - |
| MDS-UPDRS-III (ON med) | 18.3 ± 9.8 | 18.1 ± 9.5 | 17.1 ± 8.5 | ns | - |
| MDRS | 138.5 ± 5.4 | 138.5 ± 5.2 | 138.5 ± 5.7 | ns | - |
| PDQ-8 (Summary Index) | 33.9 ± 41.4 | 27.5 ± 33.1 | 27.2 ± 15.3 | ns | - |
| Dyskinesias (% yes) | 340/430 (79.1%) | 96/109 (88.1%) | 55/76 (72.4%) | **0.02** | **b>a,c** |
| On-off phenomenon (% yes) | 253/429 (59%) | 75/109 (68.8%) | 50/76 (65.8%) | ns | - |
| Wearing-off (% yes) | 393/430 (91.4%) | 104/109 (95.4%) | 73/76 (96.1%) | ns | - |
| Freezing of gait (% yes) | 209/429 (48.7%) | 51/108 (47.2%) | 23/76 (30.3%) | **0.01** | **c<a,b** |
| Orthostatic hypotension symptoms (% yes) | 34/410 (8.3%) | 17/107 (15.7%) | 10/76 (13.2%) | **0.4** | **a<b** |
| ICD (% yes) | 116/429 (27%) | 19/109 (17.4%) | 13/76 (17.1%) | ns | - |
| REM Behavior Disorders (% yes) | 168/407 (41.3%) | 45/109 (41.3%) | 42/76 (55.3%) | ns | - |
| MCI (% yes) | 38/426 (8.9%) | 9/108 (11.6%) | 10/75 (13.5%) | ns | - |

Chi-squared, logistic regression and Kruskal-Wallis non-parametric tests (with Bonferroni post hoc correction) were performed to compare demographics and clinical features at baseline between groups at a significance level of 0.05. Numerical variables are presented as means (± SD), while categorical variables are reported as absolute numbers and the percentage of “yes” responses. Abbreviations: OFF med, OFF medication; ON med, ON medication; MDRS, Mattis Dementia Rating Scale; PDQ-8, Parkinson’s Disease Questionnaire-8; ICD, impulsive-compulsive disorders; MCI, Mild cognitive impairment. Significant p-values (p<0.05) are reported in bold; ns = non-significant.

### Long-term outcome

Comparison of clinical features and long-term complications across all groups at different time points is shown in Fig. 1 and 2 and in Table 2.

**Figure 1.**
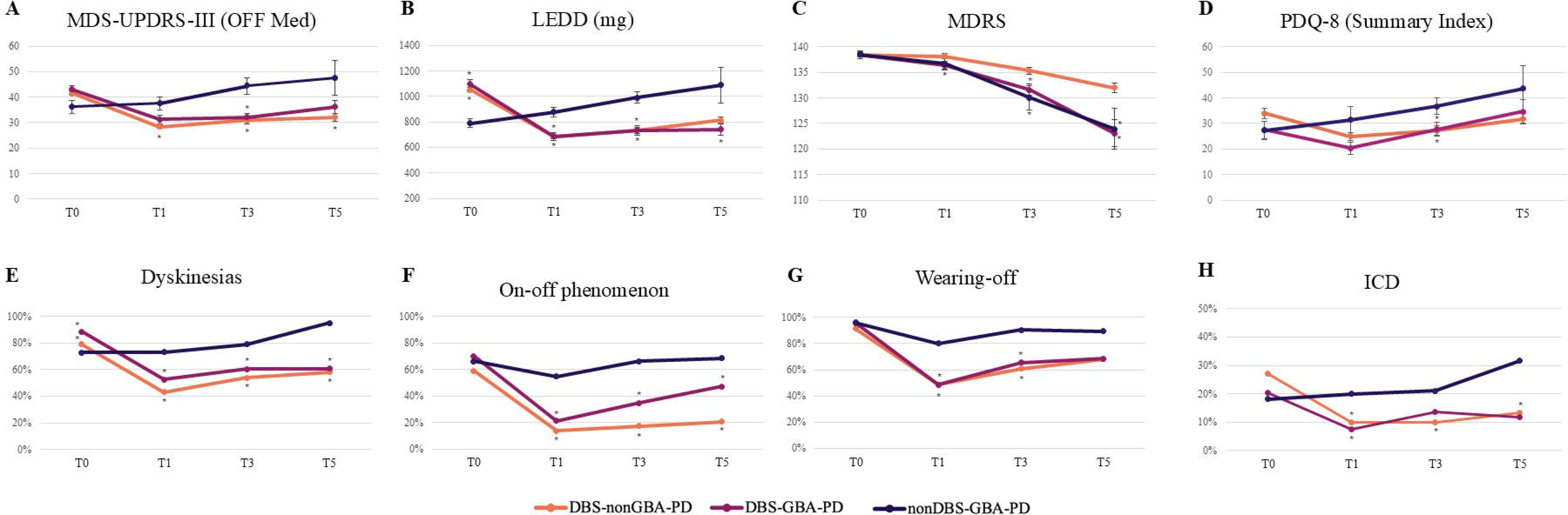
Evolution of clinical motor and non-motor parameters of the three groups over time. Graphs show mean values of (**A**) MDS-UPDRS-OFF med, (**B**) MDRS, (**C**) LEDD and (**D**) PDQ8-SI, and the percentage of (**E**) Dyskinesias, (F) On-off phenomenon, (**G**) Wearing-off, (H) ICD, at baseline (T0), T1, T3 and T5. (*) indicate statistically significant p-values (p < 0.05).

**Figure 2.**
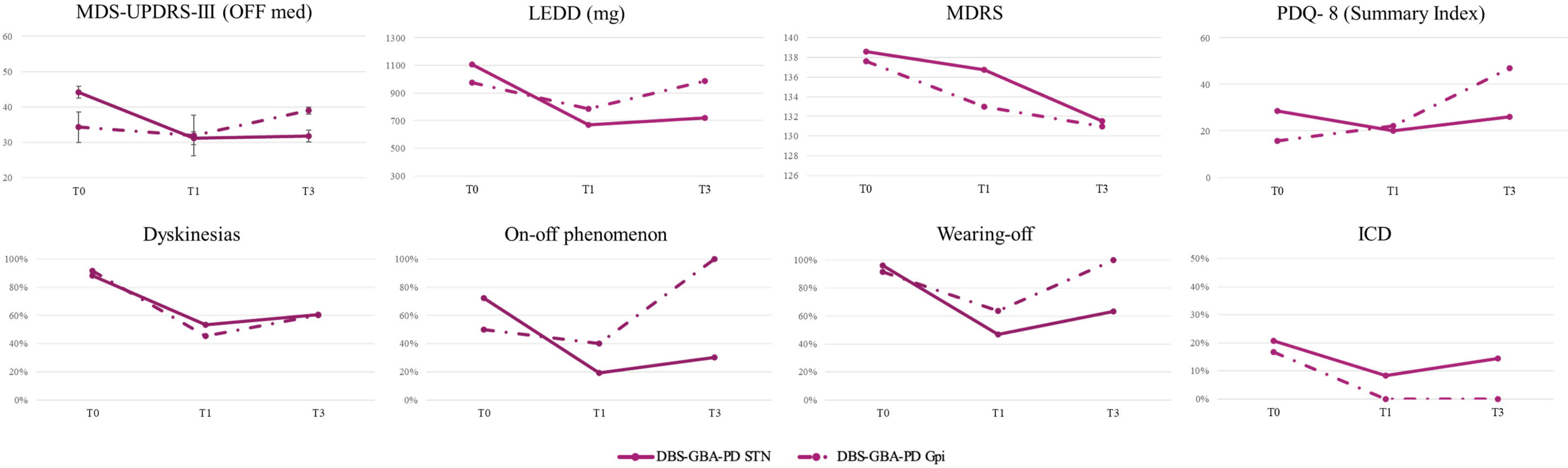
Occurrence of long-term complications in the three groups over time. Graphs show percentage of dementia, depression and hallucinations, inability to walk, recurrent falls, and urinary incontinence at T1, T3, and T5. Black (*) indicate statistically significant differences between groups at different time points; red (*) indicate significant intra-group differences over time (p < 0.05).

**Table 2.**
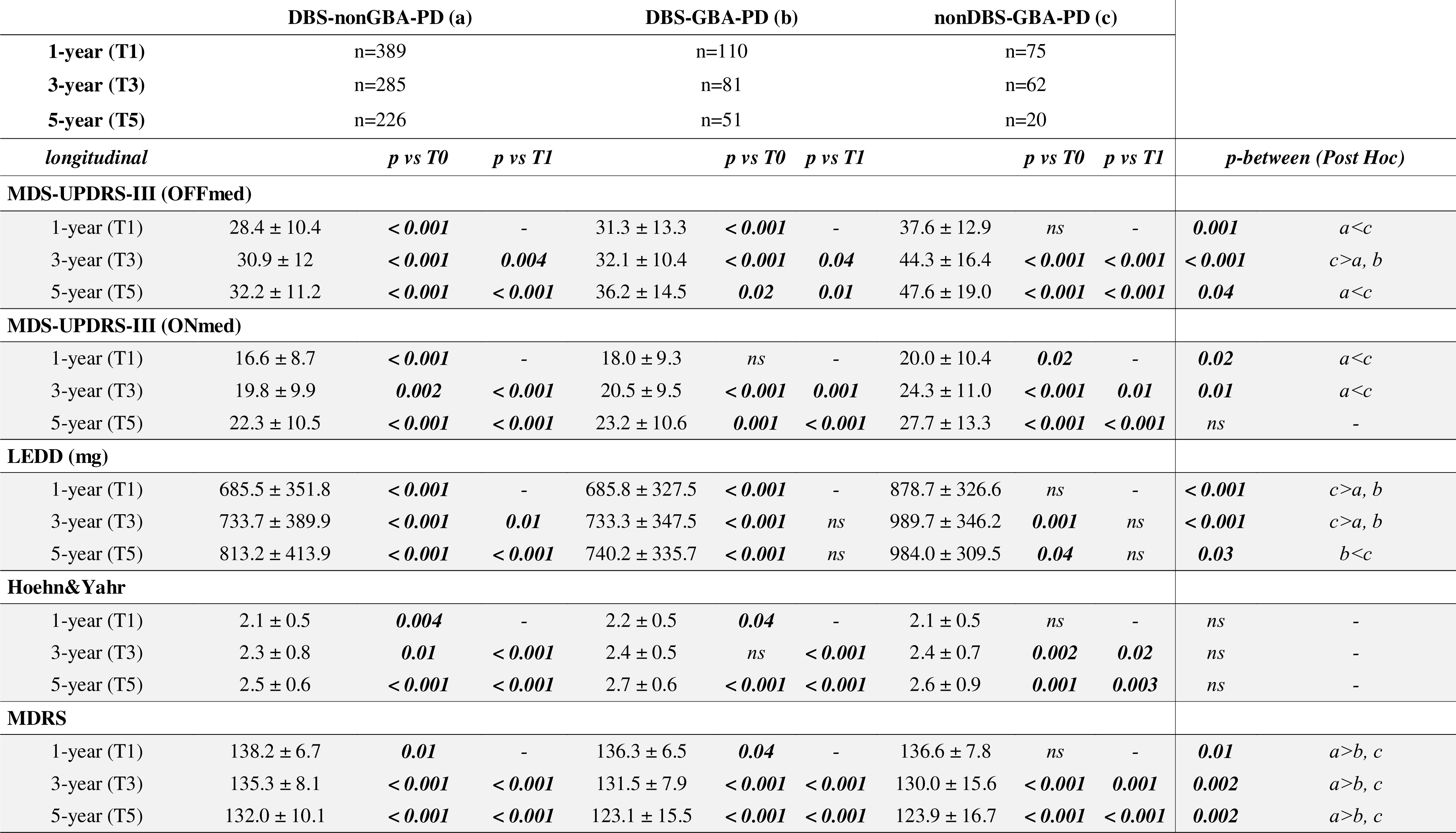
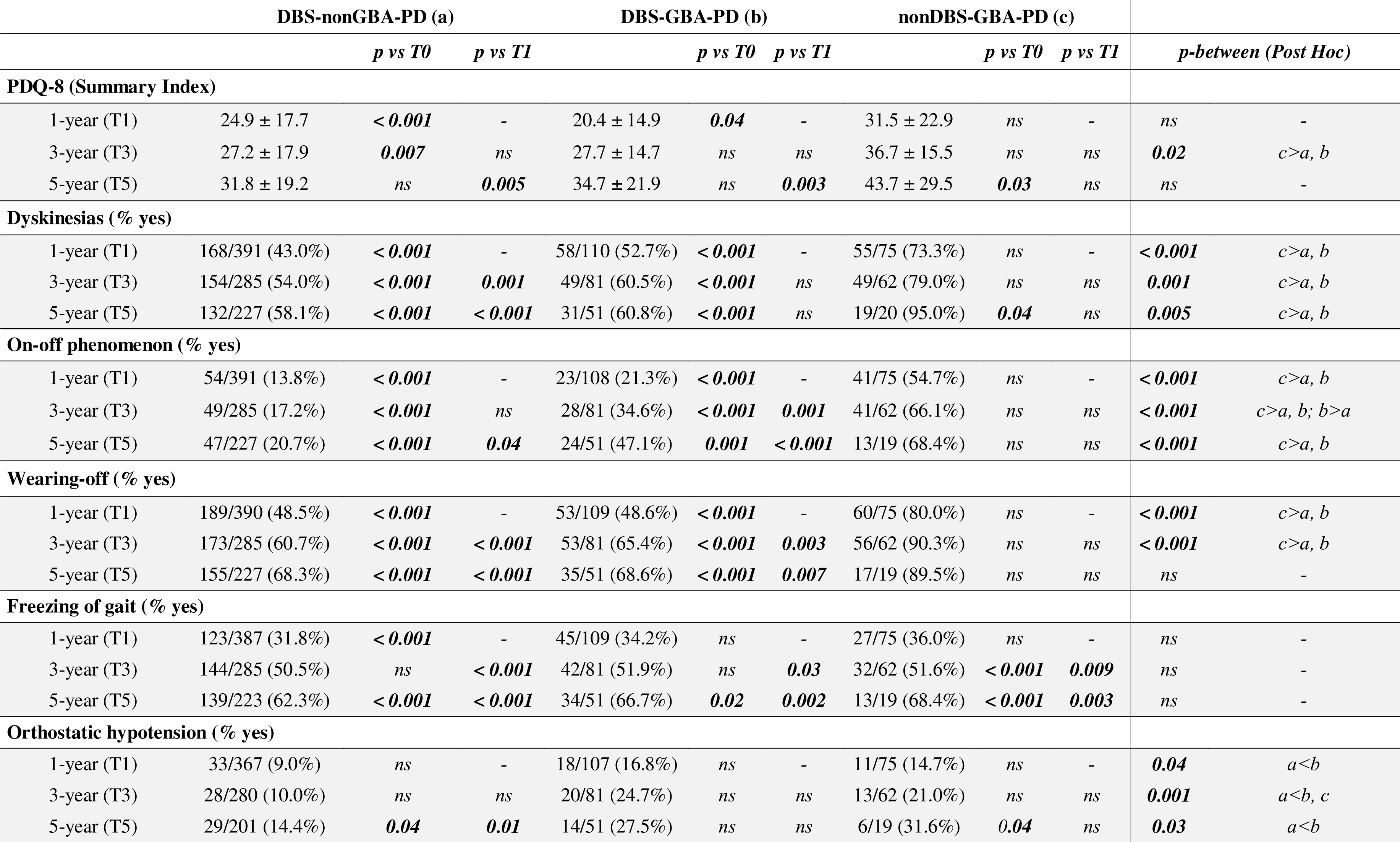
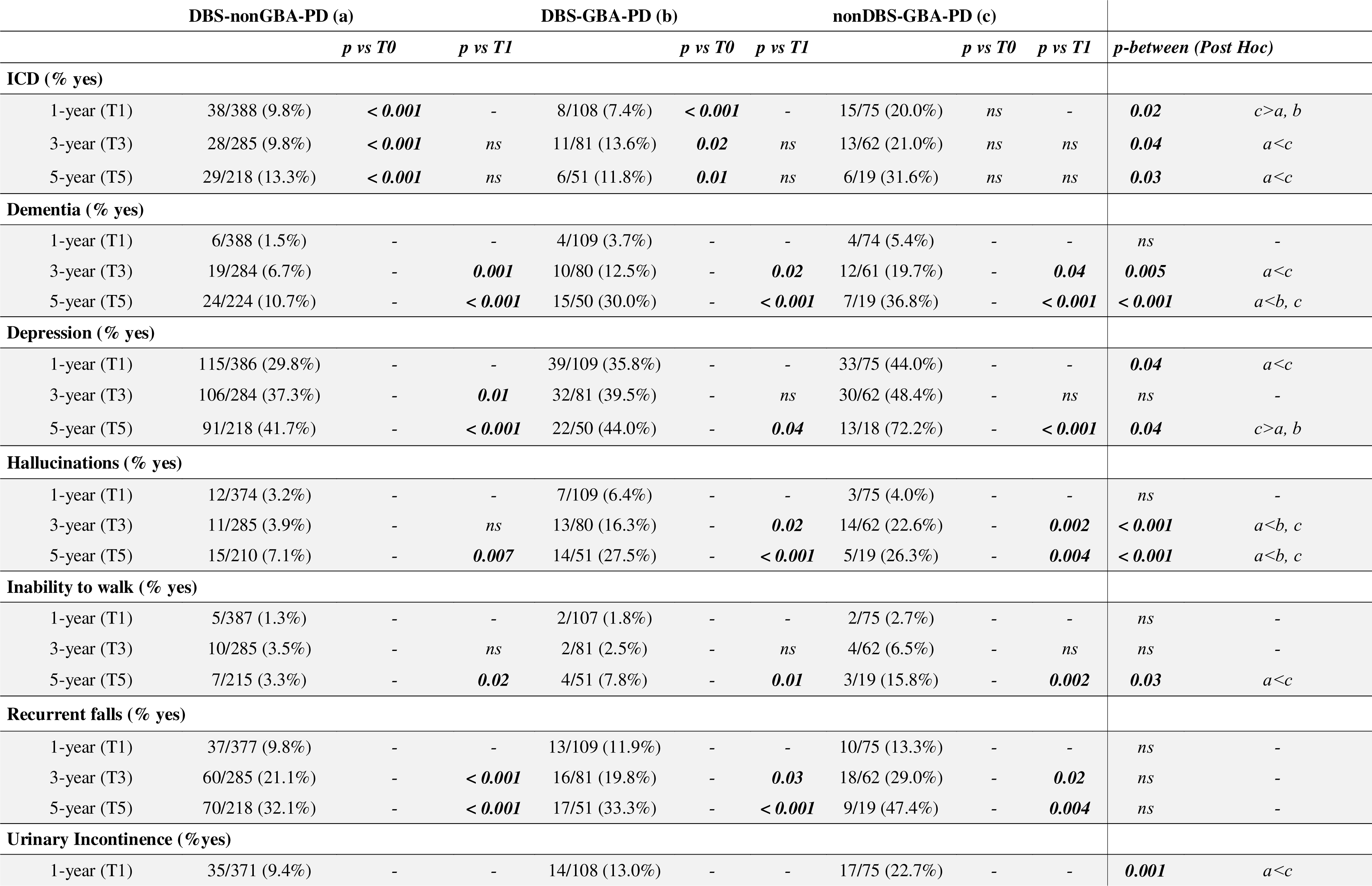
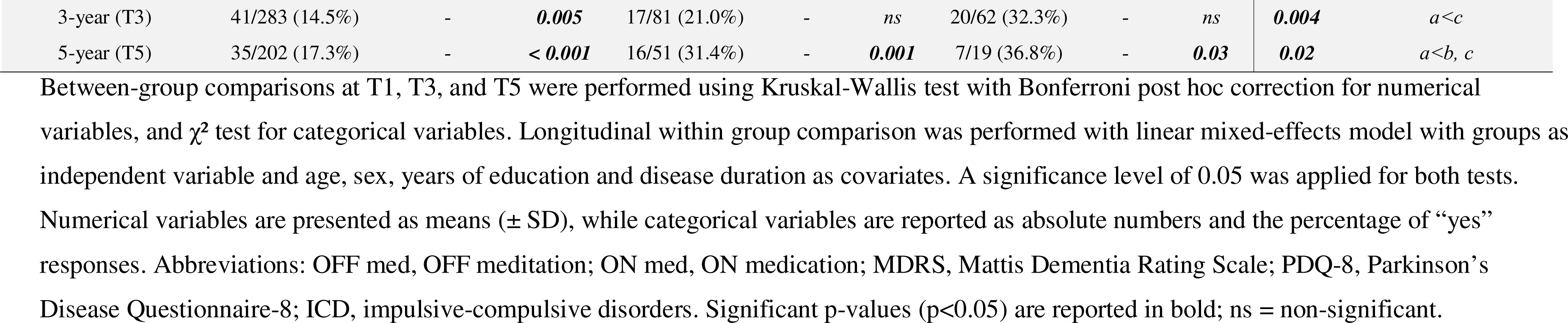
Clinical motor and non-motor parameters of the three groups at T1, T3 and T5.

At 1-year from baseline, both DBS groups, regardless of *GBA1* genotype, showed a marked improvement of motor features (decreased MDS-UPDRS-III-OFF) and motor fluctuations (reduction of dyskinesias, wearing-off, on-off phenomenon), as well as a reduction of LEDD intake and impulsive-compulsive disorders, which remained stable at the long-term follow-up. Conversely, a significant and progressive worsening in motor scores, increased LEDD intake and dyskinesias were observed in nonDBS-GBA-PD. Complications such as recurrent falls and inability to walk were limited and comparable between DBS groups, while these were more prevalent and significantly worsened from baseline in nonDBS-GBA-PD.

When assessing cognition, MDRS scores of both GBA-PD groups, regardless of DBS, showed significant worsening from baseline, differing from the DBS-nonGBA-PD group already after 1 year. At 3- and 5-years follow-up, the proportion of patients with a diagnosis of dementia was comparable in the two GBA-PD groups and significantly greater than DBS-nonGBA-PD. Similarly to cognitive decline, the prevalence of hallucinations and urinary problems also increased at 3 and 5 years in both GBA-PD groups. Depression slightly increased after 5 years in all three groups, with a significantly higher prevalence in nonDBS-GBA-PD compared to both operated groups.

Longitudinal evaluation showed that QoL significantly improved after surgery in both operated groups. Such improvement was sustained over time in DBS-nonGBA-PD, while it returned to baseline levels after 3 years and then it remained stable in DBS-GBA-PD. Conversely, QoL significantly worsened from baseline to the 5-years follow-up in the nonDBS-GBA-PD group.

### Comparison across *GBA1* variant classes

Among 109 DBS-GBA-PD subjects, mild variants were detected in 23 (21.1%) subjects, risk variants in 26 (22.9%), severe/complex variants in 47 (43.1%), and unknown variants in 13 (11.9%). Of the 76 nonDBS-GBA-PD subjects, 14 (18.4%) had mild variants, 23 (30.3%) risk, 30 (39.5%) severe/complex and 9 (11.8%) unknown (Supplementary Table 2). *GBA1* variants distribution across classes was comparable between DBS and nonDBS groups (Supplementary Fig. 1).

Demographic data at baseline are shown in Supplementary Table 3, while comparison of the longitudinal data stratified by *GBA1* variant classes up to 3-years follow-up, is shown in Fig. 3 and Supplementary Table 4.

**Figure 3.**
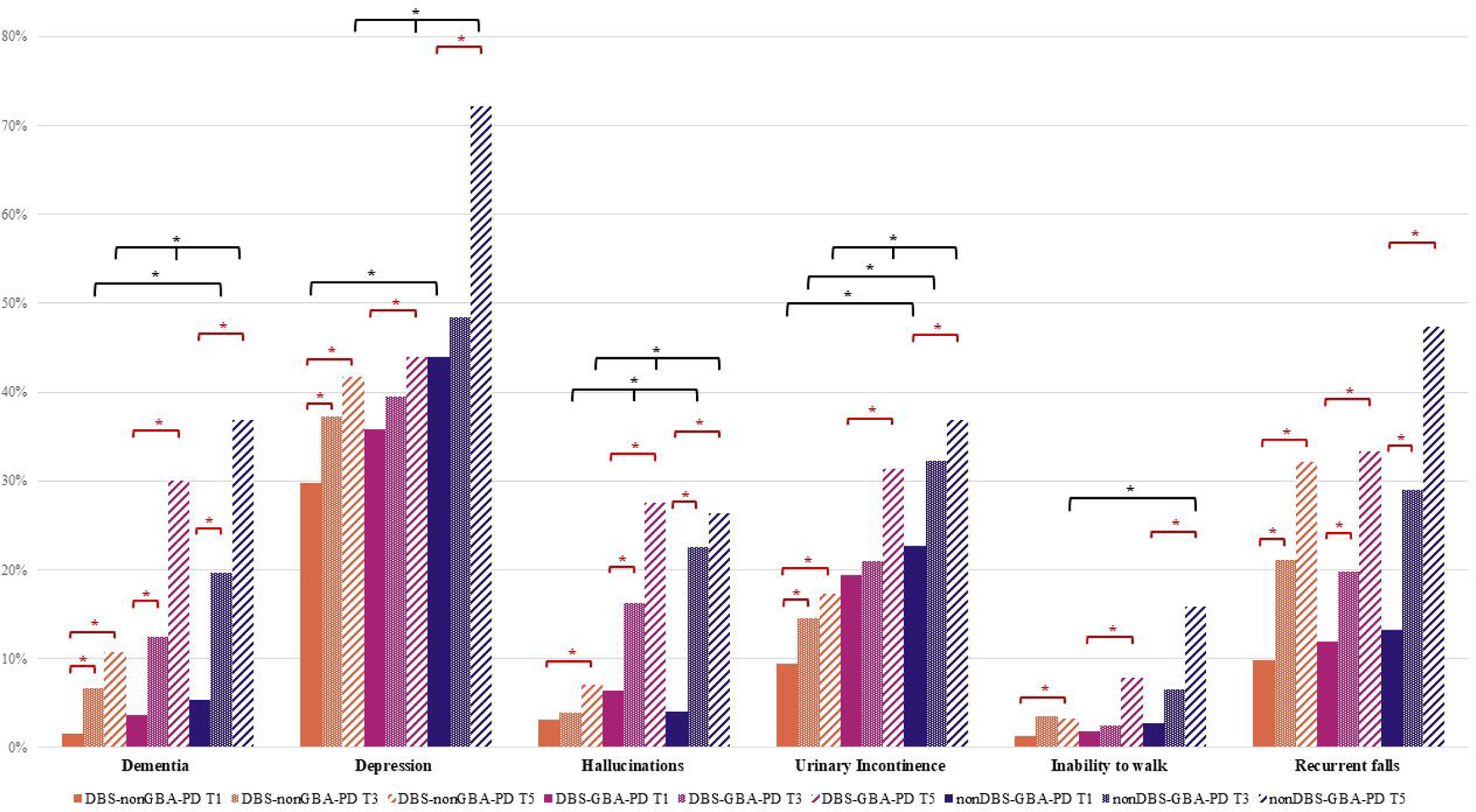
Evolution of clinical motor and non-motor parameters of GBA-PD subgroups according to variant classes. Graphs show mean values of (A) MDS-UPDRS-OFF med, (B) LEDD, (C) MDRS and (D) PDQ8-SI and the percentage of (E) Dyskinesias, (F) On-off phenomenon, (G) Wearing-off, (H) ICD, at baseline (T0), T1, and T3. Unknown variants are not shown. (*) indicate statistically significant p-values (p < 0.05).

No major differences emerged across variant subgroups for demographic characteristics, motor and non-motor features at baseline, with the exception of orthostatic hypotension symptoms, which were considerably less common in nonDBS-GBA-PD carrying risk and unknown variants compared to other subgroups. T1 and T3 evaluations also revealed a similar outcome across variant classes except for orthostatic symptoms, which were less prevalent in nonDBS-GBA-PD carrying risk variants compared to those carrying mild and severe variants. Overall, all DBS-GBA-PD variant subgroups had a comparable, prolonged benefit after surgery, with significantly reduced motor symptoms, complications and a lower LEDD. Conversely, a global motor worsening was observed in all nonDBS-GBA-PD variant subgroups.

The deterioration in MDRS scores was similar in all DBS-GBA-PD and nonDBS-GBA-PD subgroups, with a trend towards a more marked worsening in the nonDBS-GBA-PD subgroup carrying severe variants. At 3 years, no differences were observed in the prevalence of dementia across *GBA1* classes in both operated and non-operated groups.

### Outcome comparison between DBS targets

In the DBS-GBA-PD group, the target was bilateral STN in 89.4% and GPi in 10.6%. The clinical outcome up to 3 years is presented in Fig. 4 and Supplementary Table 5.

**Figure 4.**
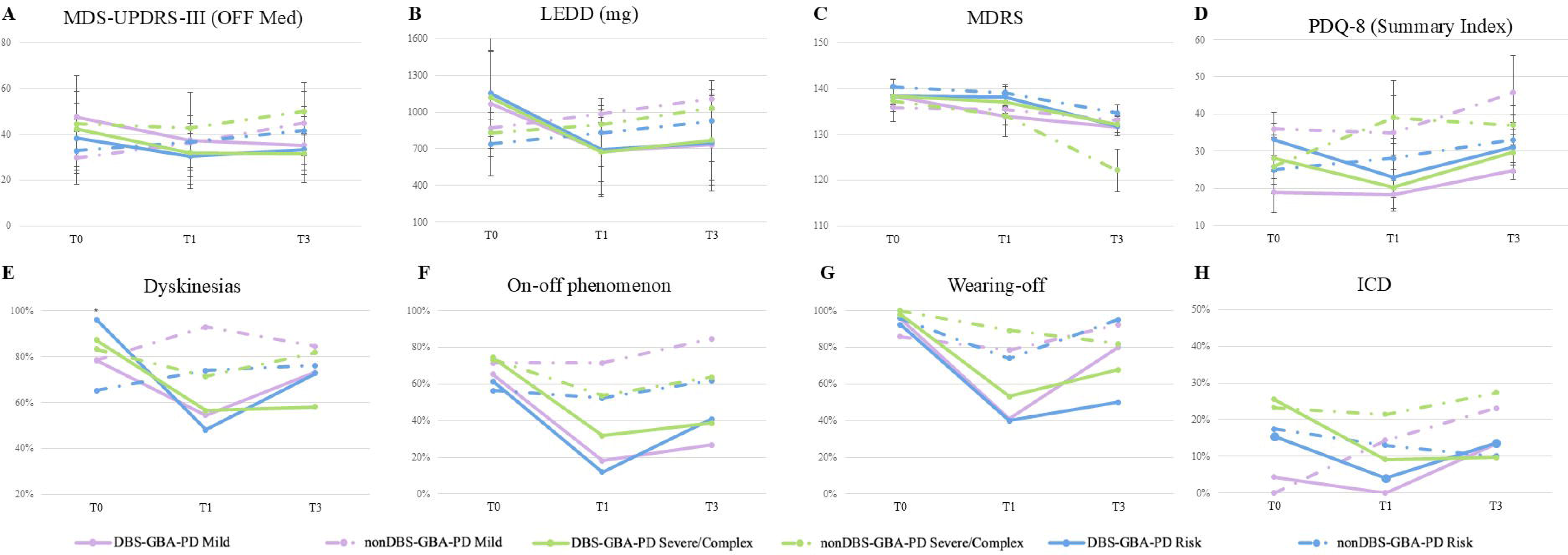
Evolution of clinical motor and non-motor parameters of DBS-GBA-PD according to DBS target. Graphs show mean values of (A) MDS-UPDRS-OFF med, (B) LEDD, (C) MDRS and (D) PDQ8-SI and the percentage of (E) Dyskinesias, (F) On-off phenomenon, (G) Wearing-off, (H) ICD, at baseline (T0), T1, and T3.

A longitudinal comparison of the estimated rate of change of motor and non-motor parameters disclosed no relevant differences between the two subgroups. Compared to baseline, we evidenced a significantly greater improvement of motor function in the DBS-GBA-PD_STN than in the GPi counterpart. To note, cognitive decline and other non-motor features evolved similarly in both subgroups regardless of the target, except for a slightly increased prevalence of hallucinations in DBS-GBA-PD_STN at the 3-year follow-up.

## Discussion

DBS is an effective treatment for advanced PD, being able to improve motor symptoms and reduce the need for medications.^25,26^ The post-surgical evolution is known to be influenced by technical aspects and clinical presentation, yet the patient’s genetic background is recently emerging as a relevant variable.^8,9,27,28^ In this light, understanding the impact of genetic risk factors on DBS response is crucial to optimise the selection of candidates and predict the clinical outcome. This is particularly relevant for *GBA1*, with a carrier frequency of ∼10% among PD.^3^ Previous studies in GBA-PD patients demonstrated that DBS is beneficial on motor symptoms and fluctuations,^10–13^ but raised concerns on its possible detrimental effects on cognitive and other neuropsychiatric symptoms.^10,12,13,29^ So far, this issue remains unresolved.

To address this key question, we expanded our former DBS cohort by including new referring centres across Italy. Before merging the newly recruited with the former cohort,^14^ a validation study was conducted to ensure that the two cohort were fully comparable in terms of the frequency of *GBA1* variant classes, baseline clinical features and post-surgical outcomes. This was an essential step, since an unbalanced distribution of *GBA1* variants or different clinical features at baseline could have biased subsequent analyses in the extended cohort. More importantly, we included a third group of GBA-PD subjects who matched the same DBS eligibility criteria as the operated cohort, but eventually did not undergo surgery. The frequency of variant classes was homogeneous in the operated and non-operated groups, with severe/complex variants being the commonest ones, in line with previous Italian studies.^14,23^

At baseline, the three cohorts were remarkably similar in terms of motor, cognitive and other non-motor symptoms such as motor fluctuations, which represented the most prevalent indication for DBS surgery. The shorter disease duration, lower LEDD and reduced freezing of gait shown by the nonDBS-GBA-PD group may be partly explained by the fact that the operated subjects were evaluated just before surgery whereas, for nonDBS subjects, the baseline was set at the time when patients were considered eligible for surgery, and thus potentially earlier than the operated groups. Evaluation up to 5 years confirmed the well described DBS-related motor improvement, which was comparable in both operated groups regardless of *GBA1* genotype, with reduced motor fluctuations and lower LEDD intake. Conversely, nonDBS-GBA-PD subjects exhibited a global decline in motor performance, which became more pronounced with time.

The most relevant finding of this study emerged when comparing the cognitive outcomes. Cognitive impairment of DBS-GBA-PD was in line with previous data.^13,14,30,31^ Non-operated GBA-PD also showed significantly lower cognitive scores than non-mutated subjects already after 1 year, and progressively worsening at longer follow-ups. Most importantly, such cognitive deterioration was remarkably similar in the two GBA-PD groups regardless of DBS, in sheer contrast with the study by Pal and collaborators, who reported a significantly worse cognitive performance in DBS-GBA-PD compared to the non-operated GBA-PD group.^13^ This discrepancy can possibly be explained by the different selection criteria adopted to recruit the nonDBS cohort. In the study by Pal *et al* ^13^, the majority of nonDBS-GBA-PD patients were in a milder stage of disease, and therefore unmatched with the operated cohort, which included patients with more advanced PD ^32^ Conversely, all non-operated GBA-PD subjects in our study fulfilled standard eligibility criteria for DBS,^21,22^ and their phenotype at baseline was largely similar to that of the two operated groups. Despite being considered appropriate candidates for DBS, eventually these patients were not operated, mainly because they preferred to delay surgery or attempt alternative therapeutic strategies.

Although some studies have implied a possible negative impact of STN-DBS on non-motor symptoms in PD cohorts untested for *GBA1*,^33–36^ others have suggested that the development of dementia may relate to the natural progression of the disease rather than the detrimental effects of DBS itself.^37–40^ By longitudinally comparing two clinically matched cohorts of operated versus non-operated GBA-PD subjects, our data supports the latter hypothesis, showing that DBS surgery does not contribute to the accelerated cognitive decline typical of the GBA-PD population.

Besides cognitive impairment, GBA-PD subjects also manifested a significantly increased burden of hallucinations compared to non-mutated cases, which was also unrelated to DBS. Our data are consistent with previous findings reporting a higher occurrence of neuropsychiatric symptoms in GBA-PD,^16,23^ suggesting that hallucinations may develop concurrently with the deterioration of cognitive functions. This observation corroborates the reported non-motor clinical similarities of GBA-PD and Dementia with Lewy Bodies (DLB),^41^ and the recent finding in GBA-PD of a significant reduction in resting activity in the posterolateral parieto-occipital cortical regions (a typical pattern of DLB), as well as the presence of a larger proportion of neocortical Lewy body pathology, compared to non-mutated cases.^42,43^

While several studies suggested that depression is more prevalent in GBA-PD,^5,16,23^ the three groups did not differ in the prevalence of depressive symptoms at baseline, likely because subjects with overt or poorly controlled depressive symptoms were excluded due to the DBS selection criteria. However, it is worth noting that, after 5 years, depression and ICD were significantly more pronounced in the nonDBS-GBA group compared to both operated groups, possibly reflecting the progressive worsening of motor function and the need for LEDD increase in patients who did not benefit from DBS treatment.^44,45^

Another poorly investigated critical issue is the long-term impact of DBS on QoL in GBA-PD. Several studies have shown an improvement of QoL soon after STN-DBS^46–48^ and up to 5-year follow-up.^49^ Only one former study evaluated QoL in a small cohort of DBS-GBA-PD patients, reporting worse scores compared to non-mutated subjects.^29^ Here we observed that, at 5 years from surgery (∼15 years from PD onset), QoL worsened significantly from baseline in non-operated GBA-PD individuals, while it remained stable or even improved in the operated groups, regardless of the *GBA1* genotype. Moreover, long-term motor complications (recurrent falls and inability to walk) were limited and comparable between DBS groups, while worsening from baseline in the non-operated group. These findings highlight how the DBS-related improvement of motor functions can positively influence QoL of GBA-PD subjects, in turn impacting healthcare costs and social burden of this common genetic condition.

A secondary objective of this study was to disclose potential differences of long-term DBS outcomes in carriers of distinct *GBA1* variant classes. Both operated and non-operated GBA-PD subjects carrying different variants classes overall exhibited a similar clinical profile, characterized by a prolonged motor benefit after surgery, and a parallel deterioration of cognitive functions and other non-motor symptoms. These observations suggest that the current classification of *GBA1* variant classes, “borrowed” from Gaucher disease,^24^ may not be appropriate to define the pathogenic impact and prognostic implication of *GBA1* variants in the context of PD, and a novel, PD-specific classification is warranted.

Finally, our exploratory analysis comparing the long-term outcome of STN vs GPi targets in GBA-PD showed similar clinical trajectories in terms of motor, cognitive and other non-motor symptoms up to 3 years post-surgery. STN was suggested to worsen cognitive deterioration and neuropsychiatric symptoms in general PD population compared to GPi;^36,50^ however, a metanalysis^51^ and a more recent study^52^ failed to detect significant differences between targets. Replication studies on larger GBA-PD cohorts comparing DBS targets are needed to confirm our observations.

We acknowledge some limitations in our study. Firstly, the three groups were not perfectly matched at baseline, as nonDBS-GBA-PD had slightly shorter disease duration, lower LEDD and reduced freezing of gait compared to operated groups. Secondly, the sample size was still relatively small when stratifying GBA-PD according to different variant classes or to different DBS targets, particularly at long-term follow-up, hampering the detection of potential subtle differences across subgroups. A third limitation resides in the retrospective nature of the study. Cognitive decline was assessed based on a single cognitive scale, unable to assess subtle differences regarding selective cognitive domains; similarly, some clinical features such as symptomatic orthostatic hypotension and urinary incontinence were mainly recorded anamnestically, possibly leading to an over-or underestimation of their frequencies.^53^

In conclusion, we report the first systematic assessment of DBS long-term impacts on GBA-PD, comparing DBS-treated subjects with a comparable cohort who did not undergo surgery. We confirm a sustained benefit of DBS on motor features, with a general improvement of QoL. Importantly, the accelerated cognitive decline of GBA-PD compared to non-mutated subjects was fully comparable in the operated and non-operated groups regardless of surgery, *GBA1* mutation class and DBS target. Overall, these results indicate that DBS should be considered as a valid and safe therapeutic option for GBA-PD.

## Data availability statement

The datasets generated during the current study are available in the ZENODO repository [DOI]:

## Acknowledgements

We are grateful to all patients for participating in the PARKNET project.

## Funding

This multicenter study was partly supported by the Italian Ministry of Health (Ricerca Corrente 2022-2024), Ricerca di Rete to the Italian Network for Neurosciences and Neurorehabilitation (RIN-RCR-2019-23669119_003 and RCR-2022-23lJ682lJ291) and National Recovery 231 and Resilience Plan (NRRP) – project MAD-2022-12376496.

## Competing interests

MA, LA was supported by Ministry of University and Research (MUR), National Recovery 231 and Resilience Plan (NRRP), project MNESYS (PE0000006).

## Appendix 1

### The Italian PARKNET Study Group

**Fondazione Santa Lucia IRCCS, Roma, Italy:** C. Caltagirone, A. Costa, F.R. Giardina, A. Peppe, C. Pellicano, F.E. Pontieri, F. Piras, F. Sommaruga, G. Sancesario, S. Zabberoni.

**Fondazione IRCCS Istituto Neurologico “Carlo Besta”, Milano, Italy:** R. Cilia, F. De Giorgi, G. Devigili, R. Eleopra, A. Elia, B. Garavaglia, G. Giaccone, M. Grisoli, V. Leta, F. Moda, B. Reggiori, L.M. Romito, I. Tramacere, S. Piacentini.

**Fondazione IRCCS Istituto Neurologico C. Mondino, Pavia, Italy:** M. Avenali, R. Bergamaschi, R. Calabrese, S. Cerri, G. Cosentino, G. Cuconato, R. De Icco, C. Galandra, A. Imarisio, P. Mitrotti, C. Pacchetti, I. Palmieri, M. Picascia, A. Pichiecchio, A. Pisani, C. Tassorelli, E.M. Valente, F. Valentino, R. Zangaglia.

**Fondazione Don Carlo Gnocchi, Milano, Italy:** C. Agliardi, F. Baglio, A. Caronni, F. Guerini, R. Mancuso, A. Mannini, M. Meloni, F. Saibene, S. Sorbi, F. Vannetti,

**IRCCS Istituto Auxologico Italiano, Milano, Italy:** A. Ciammola, L. Maderna, B. Poletti, A. Ratti, V. Silani, N. Ticozzi, F. Triulzi, F. Verde, G. Zebellin.

**IRCCS Ospedale San Raffaele, Milano, Italy:** F. Agosta, S. Amadio, V. Broccoli, R. Cardamone, M. Filippi, S. Galantucci, L. Gianolli, M. Morelli, E. Sarasso, J. Sassone, M.R. Terreni, M. A. Volontè

**Istituti Clinici Scientifici Maugeri, Pavia, Italy:** R. Bellazzi, E. Brigonzi, M. Buonocore, R. Campini, F. Cossa, E. Federici, M. Gennuso, E. Losavio, A. Losurdo, C. Lunetta, G. Maggioni, A. Malovini, C. Morasso, M. Nolano, E. Parati, C. Pistarini, D.M. Rossi, M. Terzaghi, V. Tibollo.

**Ospedale Policlinico San Martino, Genova, Italy:** L. Avanzino, G. Bonanno, L. Boni, C. Campi, E. Capello, G. Ferrara, A. Gaudio, G. Lagravinese, P. Mandich, R. Marchese, F. Massa, S. Morbelli, F. Nobili, P. Origone, E. Pelosin, L. Roccatagliata, L. Trevisan.

**IRCCS Neuromed - Istituto Neurologico Mediterraneo, Pozzilli (IS), Italy:** A. Arcella, G. Battaglia, D. Belvisi, A. Berardelli, D. Centonze, R. Ferese, S. Gambardella, A. Gialluisi, F. Giangaspero, C. Limatola, N. Modugno, P. Pantano, L. Pavone, S. Puglisi Allegra.

**IRCCS - Centro Neurolesi Bonino Pulejo, Messina, Italy:** A. Brigandì, G. Di Lorenzo, V. Lo Buono, S. Marino, S. Silvestro, C. Sorbera.

**IRCCS San Raffaele, Roma, Italy:** S. Bonassi, F. Brancati, M.F. De Pandis, B. Picconi, P.M. Rossini, F. Stocchi, L. Vacca

**IRCCS Humanitas Research Hospital, Rozzano (MI), Italy:** A. Albanese, L. Antunovic, A. Cocco, T. De Santis, R. Gatti, S. Lalli, E. Lauranzano, M. Matteoli, R. Mineri, E. Paraboschi, E. Perdixi, L. Politi, P. Polverino, D. Pozzi, G. Savini.

**Fondazione Policlinico Universitario A. Gemelli IRCCS, Roma, Italy:** Anna Rita Bentivoglio, Francesco Bove, Paolo Calabresi, Angelo Cimmino, Alessandro De Biase, Giulia Di Lazzaro, Daniela Di Giuda, Danilo Genovese, Maria Rita Lo Monaco, Martina Petracca, Carla Piano, Danilo Tiziano, Maria Gabriella Vita.

**Ospedale San Camillo IRCCS, Venezia, Italy:** G. Arcara, R. Barresi, V. Camparini, P. Cudia, G. Ferrazzi, K. Koutsikos, N. Manzo, F. Pellizzari.

**IRCCS Fondazione Ca’ Granda Ospedale Maggiore Policlinico, Milano, Italy:** A. Arighi, F. Blandini, F. Cogiamanian, M. Castellani, F. Colucci, M. Cribiù, A. Di Fonzo, G. Franco, E. Frattini, C. Losa, F. Mameli, E. Monfrini, E. Scola, I. Trezzi.

**IRCCS Istituto delle Scienze Neurologiche, Bologna, Italy:** M.G. Bacalini, L. Baldelli, G. Calandra-Buonaura, I. Cani, L. Caporali, V. Carelli, P. Cortelli, P. Ghedini, P. Guaraldi, G. Giannini, G. Lopane, R. Minardi, A. Fiorentino, F. Nonino, R. Pantieri, P. Parchi, F. Provini, C. Tonon, L. Sambati.

## Notes

### Author Declarations

Ethics committee of Full IRCCS Mondino, Pavia (IT) gave ethical approval for this work

